# Safety and Effectiveness of COVID-19 SPUTNIK V Vaccine in Dialysis Patients

**DOI:** 10.1101/2021.10.21.21265349

**Authors:** Guillermo Rosa-Diez, María Marina Papaginovic Leiva, Fernando Lombi, María Soledad Crucelegui, Ricardo Dulio Martínez, Hernán Trimarchi, Rubén Schiavelli, Mercedes Grizzo, Miguel Raño, Ricardo M. Heguilén, Rocío Anahí Jones, Luciana Gonzalez Paganti, Matías Ferrrari, Dra. Paula Zingoni, Victoria Kjohede, Jorge Raúl Geffner, Daniel Ferrante, Fernán González Bernaldo de Quirós, Vanina Pagotto

## Abstract

Given the vulnerability of people with chronic kidney disease to COVID-19, nephrology societies have issued statements calling for prioritization of these patients for vaccination. It is not yet known whether COVID-19 vaccines confer the same high level of protection in patients with kidney disease. The aims of this study were to evaluate the safety measured by the events supposedly attributed to vaccines and the effectiveness evaluated by the presence of antibodies in dialysis patients immunized with the COVID-19 Sputnik V vaccine.

**Methods:** multicenter, observational and analytical study of a prospective cohort of hemodialysis patients in the Autonomous City of Buenos Aires with a vaccination plan. Patients older than 18 years on dialysis who received both components of the COVID-19 vaccine were included.

**Results:** 491 patients included in the safety analysis. ESAVI with either the first or second component was detected in 186 (37.9% 95% CI 33.6%-42.34%). The effectiveness analysis measures of antibodies against SARS-Cov-2 were performed in 102 patients, 98% had positive IgG against SARS-Cov-2 antibodies 21 days after the second component .In patients with COVID-19 prior to vaccination, antibodies at day 21 after the first component reached almost the highest levels compared to those patients who did not have COVID-19, and the rise between the last measures was lower than patients without COVID-19.

**Conclusion:** Dialysis patients constitute a vulnerable population for SARS-Cov-2 infection, beyond the recommendations that were implemented by dialysis units, full vaccination is a priority and necessary. The Sputnik V vaccine has been shown to be safe and effective in this patient population.

## Introduction

The World Health Organization (WHO) (1) recommends vaccination against coronavirus disease 19 (COVID-19) as a fundamental primary prevention tool to limit the health and economic effects of the pandemic. As a consequence, having effective and safe vaccines in the short term, which can be used in a national strategy, will help to reduce the incidence of illness, hospitalizations and deaths related to COVID-19 and help to gradually reestablish a new normality in the functioning of our country.

On December 23, the National Administration of Medicines, Food and Medical Technology (ANMAT) submitted the report on the Sputnik V vaccine to the Ministry of Health of the Nation to advance in the Emergency Authorization of a batch series of Sputnik V vaccine, which according to preliminary phase 1/2 and 3 studies confer immunogenicity (2,3).

After the emergency authorization of the Sputnik V vaccine, other vaccines were approved for use, including the recombinant ChAdOx1-S vaccine from AstraZeneca.(4,5) and Sinopharm vaccine (4). Whilst all vaccines demonstrated efficacy in their publications (2–4,6), they come from phase 3 studies, which evaluate the efficacy of the vaccine. Effectiveness refers to the protection provided by the vaccine as measured in observational studies that include people with underlying medical conditions who have been receiving vaccines from different healthcare providers under real-world conditions.

So far there are no observational epidemiological studies that reveal the behavior of those vaccinated for COVID-19 in Argentina and therefore evaluating their effectiveness and reactogenicity. The safety profile of vaccines and reactogenicity are fundamental elements for the acceptance of vaccines in the population. If a vaccine is perceived as too reactogenic, the subject may refuse additional doses or the healthcare professional may choose not to recommend it, which can lead to incomplete protection of the individual and low coverage of the vaccine in the population (7,8).

Maintaining high vaccine coverage is critical to the success of vaccination programs. A few years ago, he was able to consider the concept “no pain, no gain”, assuming that, if a vaccine does not produce inflammation as a “proxy” for pain, the immune response achieved was poor, leaving a common belief that a reaction in place of injection to a vaccine is a predictive sign of a desirable vaccine response (9).

The Ministry of Health of the Nation (10), has designed the Strategic Plan for vaccination against COVID-19 in Argentina (11), being one of its specific objectives being establish essential priority axes to evaluate vaccination goals: coverage rates, continuous monitoring of safety and effectiveness of vaccines, in correlation with the epidemiological impact that vaccination produces on COVID-19 *(11)*.

Kidney disease substantially increases the risk of severe COVID-19. Considering the relative efficacy of the current therapeutic arsenal available to reduce hospitalizations and mortality from COVID-19, and effective and safe vaccination are currently the only realistic options to curb the ongoing pandemic and reduce SARS CoV-2 infections. Throughout 2020, several vaccines were licensed for emergency use, and many more are in development (12).

Dialysis patients constitute a population at risk, not only because of their vulnerability to COVID-19, but because they cannot comply with social isolation since they must go to dialysis every three weeks. The COVID Registry of the Argentine Dialysis Registry showed that COVID infection affected 12% of the population on dialysis to date, and that mortality was 11 times higher compared to the general population (13).

Given the vulnerability of people with chronic kidney disease (CKD) to COVID-19, leading nephrology societies such as the UK Kidney Association and the US National Kidney Foundation have issued statements calling for prioritization of these patients for vaccination. The Argentine Society of Nephrology presented it in a timely manner to the Ministry of Health of the Nation, which has considered the priority of this patient population. However, it is not yet known whether COVID-19 vaccines confer the same high level of protection in patients with kidney disease as reported for participants in recent trials, who were generally healthy (14).

Patient characteristics such as age, sex, type of kidney disease, and treatment regimen can also influence the inmunological response to vaccines. In patients with weak or no induction of T-cell seroconversion and / or immunity after vaccination, theoretical options include an additional booster dose, administration of a different vaccine platform (15)

Seroconversion after confirmed infection would approach 100% in the dialysis population, but the durability of this immune response and the degree to which it translates into protective immunity remain unclear. Some studies indicate that SARS-CoV-2 IgG titers decrease substantially 3 months after diagnosis (16). Ongoing research should elucidate whether certain vaccines offer specific benefits for people on chronic dialysis. The aims of this study were to evaluate the safety measured by the events supposedly attributed to vaccines and the effectiveness evaluated by the presence of antibodies in dialysis patients immunized with the COVID-19 Sputnik V vaccine.

## Materials and Methods

### Ethics statement

The study was approved by the Ethics Committee of the Alberto C Taquini Institute for Translational Medicine Research of the Faculty of Medicine of the University of Buenos Aires. For the effectiveness study, based on the measurement of antibodies, informed consent was requested. The study protocol was registered in ClinicalTrials.gov Identifier: NCT04944433

### Population and settings

A multicenter, observational and analytical study was carried out on a prospective cohort of hemodialysis patients in the Autonomous City of Buenos Aires with a vaccination plan. The date of inclusion in the cohort was the start date of vaccination. Patients older than 18 years on dialysis who received both components of the COVID-19 vaccine were included.

Patients of the following the dialysis centers were included:Cemic Saavedra; Centro Integral de Nefrología; Centro de Diálisis Lacroze; Centro Médico FINAER; ; Clínica San Camilo; Dialitys S.A; Diaverum Argentina S.A, sede Barracas; Diaverum Argentina S.A., sede Palermo; Diaverum Argentina S.A, sede Paternal, Diaverum; FME Avellaneda; FME Caballito; FME Ciudad Evita; FME Ciudadela; FME Mansilla; FME Martínez; FME Morón; FME San Fernando; FME San Justo; Fundación Favaloro; Hospital Británico; Hospital Italiano de Buenos Aires; Hospital Donación Francisco Santojanni; Hospital General de Agudos Dr. Juan A. Fernández; Hospital Militar Central Cirujano Mayor Dr. Cosme Argerich; Hospital General de Agudos Carlos G. Durand; Hospital General de Agudos Dr. Cosme Argerich; Hospital Naval Dr. Pedro Mallo; Hospital Médico Policial Churruca Visca; Hospital Aeronáutico Central; Hospital de Clínicas José de San Martín; IART; Instituto de Nefrología del Oeste S.R.L.; Instituto Renal Metropolitano S.A; Servicio de Terapia Renal Argentina S.A sucursal Flores; Servicio de Terapia Renal Argentina S.A sucursal Pringles.

### Data collection and analysis methods

Data related to safety were collected, events allegedly attributed to vaccines and immunizations (ESAVI) after receiving the Sputnik V vaccine, the history of COVID-19 prior to vaccination and the presence of symptomatic COVID-19 after it. Demographic and kidney disease-related (type of dialysis: hemodialysis or peritoneal; kidney transplant; time in dialysis) data were obtained This information was collected by the health professionals of each center who are in charge of the dialysis sessions of the patients using an epidemiological file designed for the study was used.

Data referring to the effectiveness was the measurement of antibodies type immunoglobulin G (IgG) against SARS-Cov-2. The “COVIDAR IgG” test, which is registered in ANMAT, was used for the determination of antibodies. The test detects in blood and serum antibodies that the immune system produces for the new coronavirus, specifically against two viral antigens: the spike protein (S) and the receptor-binding domain (RBD). It is performed on plates that allow testing 96 sera at the same time using the technique known as ELISA, the same one used, for example, for the detection of HIV infection and hepatitis B. The COVIDAR IgG test detects the presence of IgG qualitatively and semi-quantitatively. In the semi-quantitative determination, the values are measured in absorbance levels (DO) with a maximum of 3.3 and a inferior limit of detection of 0.3.The processing of the samples and the performance of the ELISA both for detection of the SARS-Cov2 was carried out by the virology laboratory of the Faculty of Medicine of the University of Buenos Aires and the measurements were made before the administration of the first component, at 21 days of the same and at 21 days of the second component.

### Sample Size and Statistical analysis

For the safety objective, considering a prevalence of ESAVI with the SPUTNIK V vaccine of 60% (17) with a precision of 5% for a confidence interval of 95%, a sample size of 369 patients was estimated.

For the effectiveness objective considering the publication of the published phase 1/2 results of the Sputnik V vaccine study(2) and assuming that the vaccinated population in Buenos Aires will have the same behavior as published, the following sample scenarios were evaluated:

a) For a delta of IgG antibody titers between 0 and 21 days of 1.24 with a standard deviation of 1 with a power of 90% and an alpha of 0.01, for a two-tailed hypothesis test. The sample size is 11 people

b) For a delta of IgG antibodies between 14 to 21 days of 0.57 with a standard deviation of 1 with a power of 90% and an alpha of 0.01, for a two-tailed hypothesis test: The sample size is 49 people.

Adjusting for a 20% loss to follow-up and considering that the immunogenicity of these patients is lower than that of the general population, the calculation of the sample size was estimated at 100.

In the descriptive analysis, quantitative data were expressed as median and interquartile range (IQR) according to their distribution. Qualitative data were expressed as absolute (n) and relative (%) frequencies. To allow external comparisons with other publications, we also defined a dichotomous classification in individuals ≤ 55 or> 55 years old. For the comparisons according to the presence of ESAVI, the T or Wilcoxon test was used for the quantitative data according to their distribution and the Chi square of Fisher test according to the assumptions.

The proportion of patients with ESAVI was estimated with its 95% confidence interval (95% CI). To evaluate the factors associated with the presence of ESAVI, a multiple logistic regression was performed, considering as independent variables those statistically significant in the bivariate analysis and those that were clinically significant according to the research team. The crude Odds ratio (OR) were expressed and adjusted with their 95% CI.

A random effect fixed model was used to compare the immunoglobulin G levels for coronavirus type 2 that causes severe acute respiratory syndrome. A level of statistical significance less than 5% was considered. The analysis was carried out with software R version 4.0.3

### Funding source

The COVIDAR group provided the Serokits for sampling and the ELISA COVIDAR IgG kits, supported by FOCEM and Asoc. SAND. None of the funding sources provided economical support for the data collection, statistical analysis, or were used to write the manuscript, or to submit it for publication.

## Results

A total of 996 patients were vaccinated with the two components of the Sputnik V vaccine and 491 were included in the safety analysis. They had at least one ESAVI with either the first or second component 186 people 37.9% (95% CI 33.6%-42.34%), with the first component 112 (28.3%), and 99 (20.2%) with the second. Antipyretic before the second component was referred by 60 patients (12.2%) Of those 99 people who had ESAVI both with the first and second component a total of 54 (54.5%) perceived that symptoms were greater with the second componente than the first, while 13 (13.1%) perceived second component had less symptoms and 32 (32.3%) equal. Table 1 shows the characteristics of the total number of patients and comparison according to the presence or absence of ESAVI with any component of the vaccine.

**Table 1.** Characteristics of the total number of patients and comparison according to the presence or absence of events supposedly attributed to vaccines and immunizations with any component of the Sputnik V vaccine.

| Characteristic | Total<br>n= 491 | ESAVI<br>n=186 | NO ESAVI<br>n=305 | p valor |
| --- | --- | --- | --- | --- |
| Female<sup>1</sup> | 194 (39.5) | 81 (43.5) | 113 (37.0) | 0.182 |
| Age in years at first component<sup>2</sup> | 54.3 (43.3-64.2) | 50.1 (38.3-60.2) | 57.1 (46.5-65.5) | <0.001 |
| >50 years<sup>1</sup> | 242 (49.3) | 71 (38.2) | 171 (56.1) | <0.001 |
| Hemodialysis<sup>1</sup> | 470 (95.7) | 177 (95.2) | 293 (96.1) | 0.802 |
| Time on dialysis in years<sup>2</sup> | 3.6 (1.8-5.8) | 3.5 (1.8-6.4) | 3.6 (1.8-5.2) | 0.284 |
| Comorbidities |  |  |  |  |
| Vaccine allergies<sup>1</sup> | 15 (3.1) | 11 (5.9) | 4 (1.3) | 0.009 |
| Diabetes<sup>1</sup> | 109 (22.2) | 32 (17.2) | 77 (25.2) | 0.049 |
| Hihg blood preasure | 299 (60.9) | 108 (58.1) | 191 (62.6) | 0.363 |
| Hepatitis C<sup>1</sup> | 10 (2.0) | 6 (3.2) | 4 (1.3) | 0.189 |
| Hepatitis B<sup>1</sup> | 0 | 0 | 0 |  |
| HIV<sup>1</sup> | 4 (0.8) | 1 (0.5) | 3 (1) | 0.999 |
| Dyslipemia<sup>1</sup> | 86 (17.5) | 25 (13.4) | 61 (20.0) | 0.083 |
| Coronary heart disease<sup>1</sup> | 30 (6.1) | 12 (6.5) | 18 (5.9) | 0.958 |
| Epilepsy<sup>1</sup> | 9 (1.8) | 4 (2.2) | 5 (1.6) | 0.736 |
| Autoimmune diseases<sup>1</sup> | 26 ( 5.3) | 13 (7.0) | 13 (4.3) | 0.271 |
| Malnutrition<sup>1</sup> | 15 ( 3.1) | 4 (2.2) | 11 (3.6) | 0.523 |
| Kidney transplant<sup>1</sup> | 59 (12.0) | 36 (19.4) | 23 (7.5) | <0.001 |
| COVID before vaccination<sup>1</sup> | 66 (13.5) | 29 (15.6) | 37 (12.2) | 0.280 |
| NSAIDs prior to vaccination<sup>1</sup> | 60 (12.2) | 22 (11.8) | 38 (12.4) | 0.948 |

<sup>1</sup>n (%) <sup>2</sup>median (IQR)

There were 355 ESAVI, because patients had more than one ESAVI. No events of special interest were observed (vaccine-augmented disease; multisystemic inflammatory syndrome; respiratory distress; acute heart failure; cardiomyopathy; arrhythmias; coronary artery disease; myocarditis; acute kidney failure; acute liver failure; Guillán Barré; encephalopathy; acute disseminated encephalomyelitis; transverse myelitis; seizures; meningoencephalitis; thromboembolism; thrombocytopenia vasculitis; acute septic arthritis; erythema multiforme; perneum erythema; anaphylaxis)

Of the total ESAVI, the most frequent was pain at the injection site with both components of the vaccine, new or worse muscular pain and fever. All ESAVI were more frequent with the first componente except pain at the injection site, which was the same in both components and vomiting, that was more frequent with the second component. Figure 1 shows the frequency of ESAVI globally and after each component.

**Figure 1.**
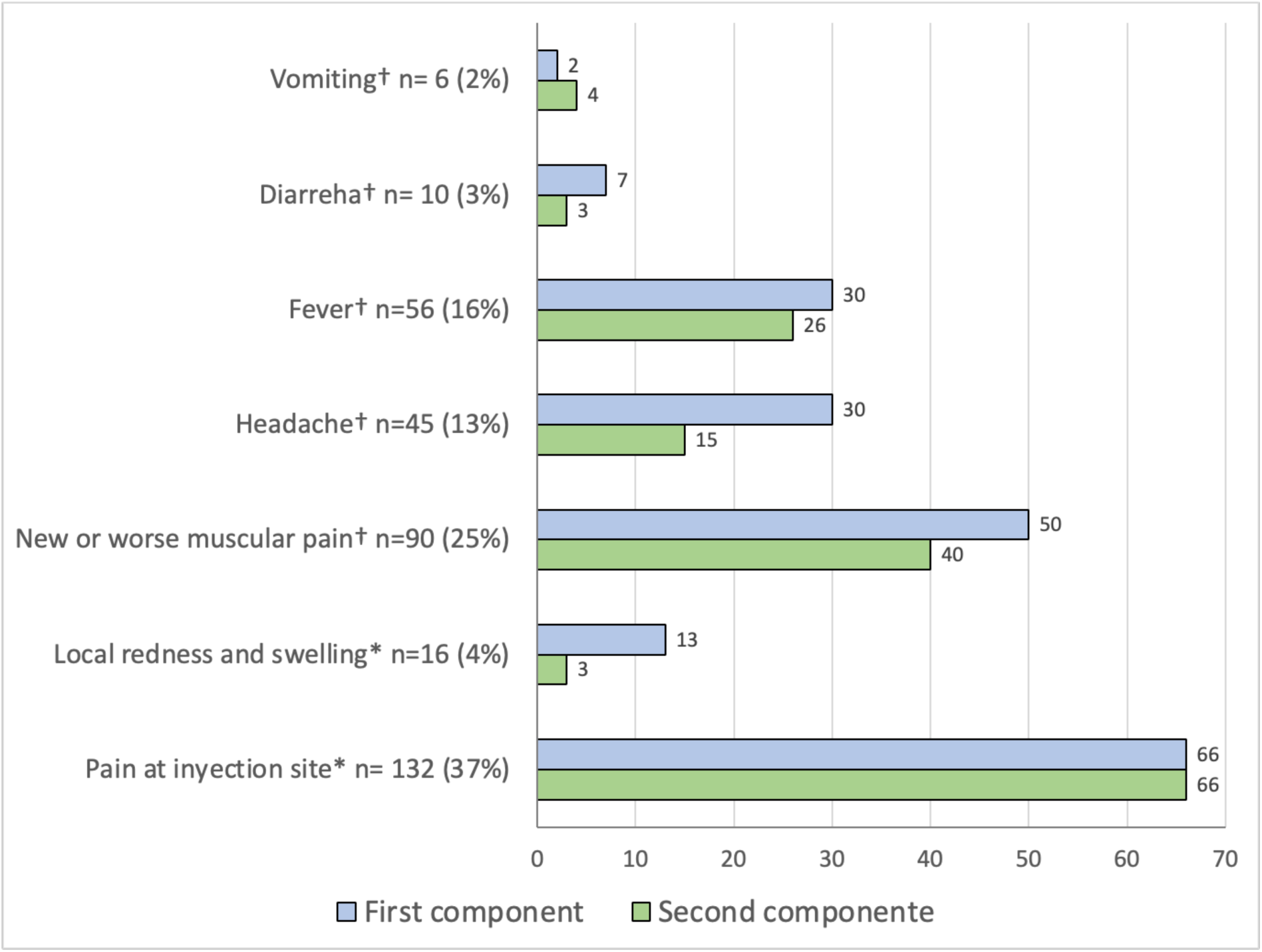
Local and systemic reactions in dialitic patients globally and after each component of Sputnik V COVID-19 vaccine. n=355 *Local reactions, †Systemic reactions

Considering having any ESAVI with either the first or the second component, Any allergy prior to vaccination, being 55 years or younger and renal transplantation were predictors of EASVI (table 2)

**Table 2.**
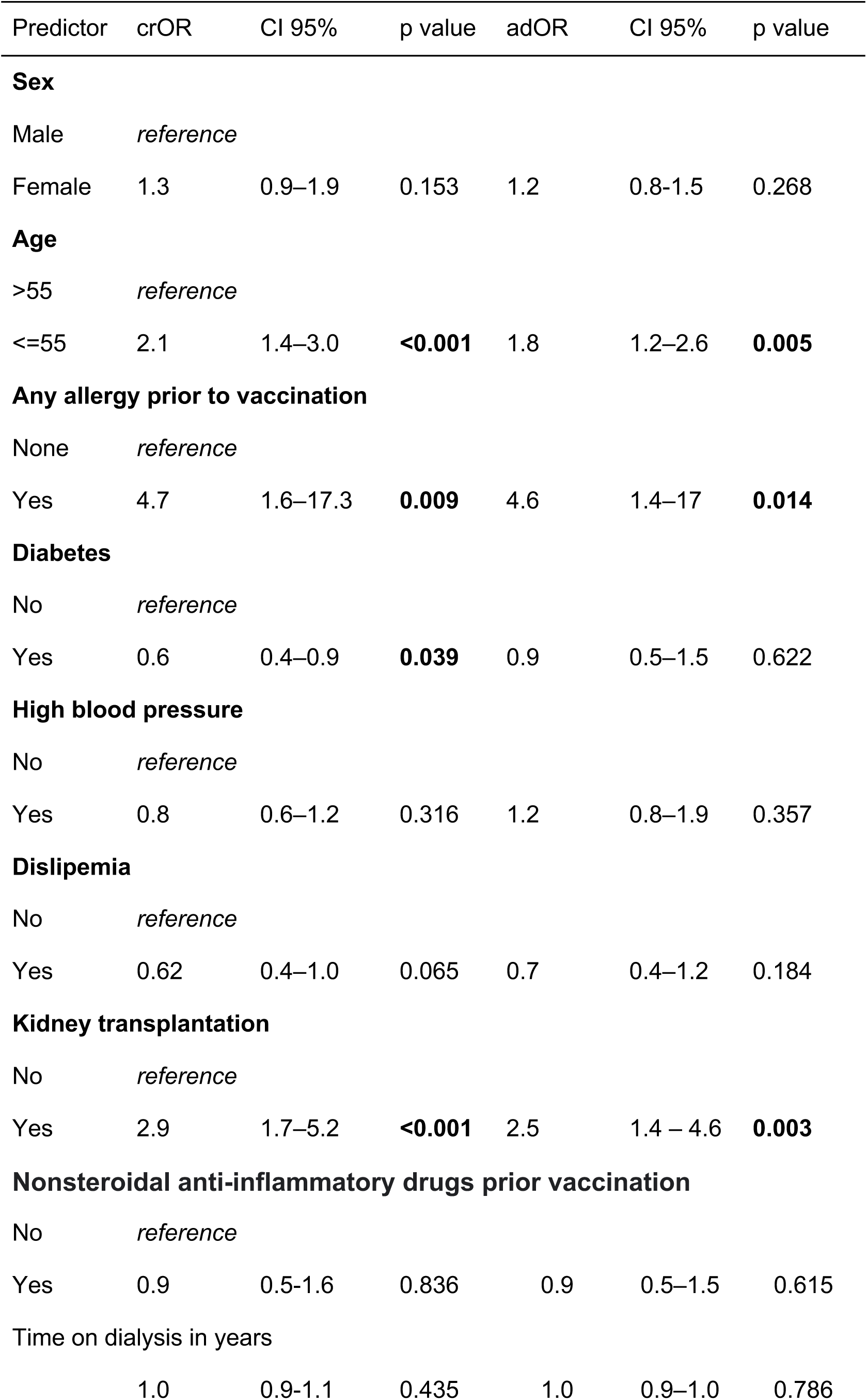
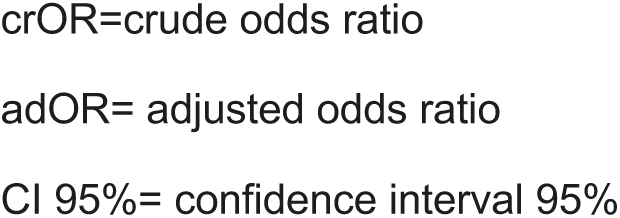
Predictor of Events Supposedly Attributed to Vaccines and Immunizations (ESAVI) in dialitic patient after Sputnik V COVID-19 vaccine

| Predictor | crOR | CI 95% | p value | adOR | CI 95% | p value |
| --- | --- | --- | --- | --- | --- | --- |
| **Sex** |  |  |  |  |  |  |
| Male | *reference* |  |  |  |  |  |
| Female | 1.3 | 0.9–1.9 | 0.153 | 1.2 | 0.8–1.5 | 0.268 |
| **Age** |  |  |  |  |  |  |
| >55 | *reference* |  |  |  |  |  |
| ≤55 | 2.1 | 1.4–3.0 | **<0.001** | 1.8 | 1.2–2.6 | **0.005** |
| **Any allergy prior to vaccination** |  |  |  |  |  |  |
| None | *reference* |  |  |  |  |  |
| Yes | 4.7 | 1.6–17.3 | **0.009** | 4.6 | 1.4–17 | **0.014** |
| **Diabetes** |  |  |  |  |  |  |
| No | *reference* |  |  |  |  |  |
| Yes | 0.6 | 0.4–0.9 | **0.039** | 0.9 | 0.5–1.5 | 0.622 |
| **High blood pressure** |  |  |  |  |  |  |
| No | *reference* |  |  |  |  |  |
| Yes | 0.8 | 0.6–1.2 | 0.316 | 1.2 | 0.8–1.9 | 0.357 |
| **Dislipemia** |  |  |  |  |  |  |
| No | *reference* |  |  |  |  |  |
| Yes | 0.62 | 0.4–1.0 | 0.065 | 0.7 | 0.4–1.2 | 0.184 |
| **Kidney transplantation** |  |  |  |  |  |  |
| No | *reference* |  |  |  |  |  |
| Yes | 2.9 | 1.7–5.2 | **<0.001** | 2.5 | 1.4 – 4.6 | **0.003** |
| **Nonsteroidal anti-inflammatory drugs prior vaccination** |  |  |  |  |  |  |
| No | *reference* |  |  |  |  |  |
| Yes | 0.9 | 0.5–1.6 | 0.836 | 0.9 | 0.5–1.5 | 0.615 |
| **Time on dialysis in years** |  |  |  |  |  |  |
|  | 1.0 | 0.9–1.1 | 0.435 | 1.0 | 0.9–1.0 | 0.786 |

crOR=crude odds ratio
adOR= adjusted odds ratio
CI 95%= confidence interval 95%

Eighteen patients had COVID-19 after the first component, fifteen were symptomatic and 17 detected by PCR; only in one patient the diagnosis was for epidemiological nexus. The median of days between the first component and COVID-19 was 23.5 days with a minimum of 2 days and a maximum of 65 days. All cases were mild.

The effectiveness analysis measures of antibodies against SARS-Cov-2 were performed in 102 patients, of whom 50 (49.0%) were female, median age was 51.6 years (IQR 39.8-62.0) and were older than 55 year 42 (41.0%). Had COVID-19 prior to vaccination 16 patients (15.7%), with a median from the diagnoses to the administration of the first component of 7.0 month (IQR 6-8). Median time for the administration of the second component were 2.8 moth (IQR 2.7-2.9) Of the 102 patient, twenty seven (26.5%) had positive IgG against SARS-Cov-2 in baseline measure, and 13 did not refer had COVID-19, which implies a rate of asymptomatic covid disease of 13/102= 12.7%. Median time in dialysis was 2.9 years (IQR 1.5-5.6).

Nine patients had COVID-19 between the first and the second component and 6 had positive antibodies measured 21 days after the first component and all had positive IgG 21 days after the second component.

Ninety eight percent of the patients had positive IgG against SARS-Cov-2 antibodies 21 days after the second component. Among the 16 patients that had COVID-19 before the first component, fourteen had positive IgG in baseline measure and only two hadn’t got positive IgG 21 days after the second componente (figure 2).

**Figure 2.**
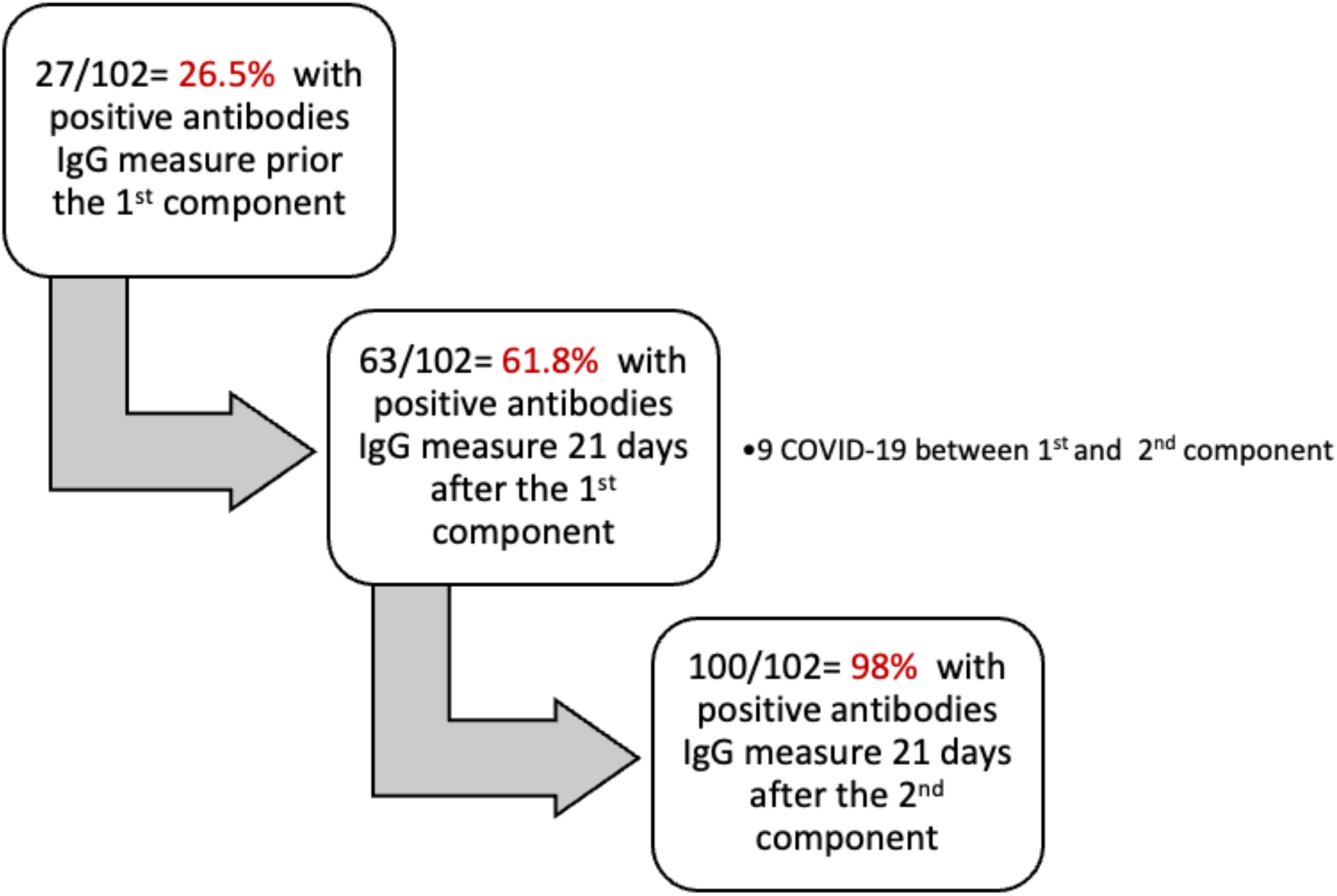
Flowchart of the antibody positivization sequence against SARS-Cov-2

Neither of these two patients, who did not show IgG at day 21, had COVID-19 before the administration of the first component of the vaccine or between the first and the second component. Both patients had hemodialysis as dialysis treatment; one was male and the other female with ages of 71 and 55 years old and one had a mild adverse reaction to vaccine and the other no, respectively.

There were differences in the levels of IgG antibodies in each time measure between patients with or without COVID-19 prior to vaccination. The first had higher levels in every measure. Patines without COVID-19 had the baseline measure under the detection limit (<0.38). Both group of patients showed a significant rise in the level of IgG en the 3 measures (table 3)

**Table 3.** Comparison of antibodies against SARS-Cov-2 in every measure according to group of the presence of COVID before vaccination and in every group time measure

| Measure | No COVID-19<br>before vaccination | COVID-19<br>before vaccination | p value<br>between<br>groups |
| --- | --- | --- | --- |
| Baseline | 0.22 (0.20-0.24) | 2.54 (2.04-2.89) | $<0.001^\dagger$ |
| 21 days after 1<sup>st</sup> component | 0.35 (0.22-1.14) | 3.37 (3.34-3.4) | $<0.001^\dagger$ |
| 21 days after 2<sup>nd</sup> component | 3.24 (1.96-3.37) | 3.38 (3.36-3.40) | $<0.001^\dagger$ |
| P value intra group | $p<0.001^*$ | $<0.001^*$ | |

$^\dagger$ wilcoxon test comparing median in each time measure by group whether had COVID=19 before vaccination or not
$^*$ random effect fixed model comparing median in 3 measures in each group whether had COVID=19 before vaccination or not

In patients with COVID-19 prior to vaccination, antibodies at day 21 after the first component reached almost the highest levels compared to those patients who did not have COVID-19, and the rise between the last measures was lower than patients without COVID-19 (table3 and figure 3).

**Figure 3.**
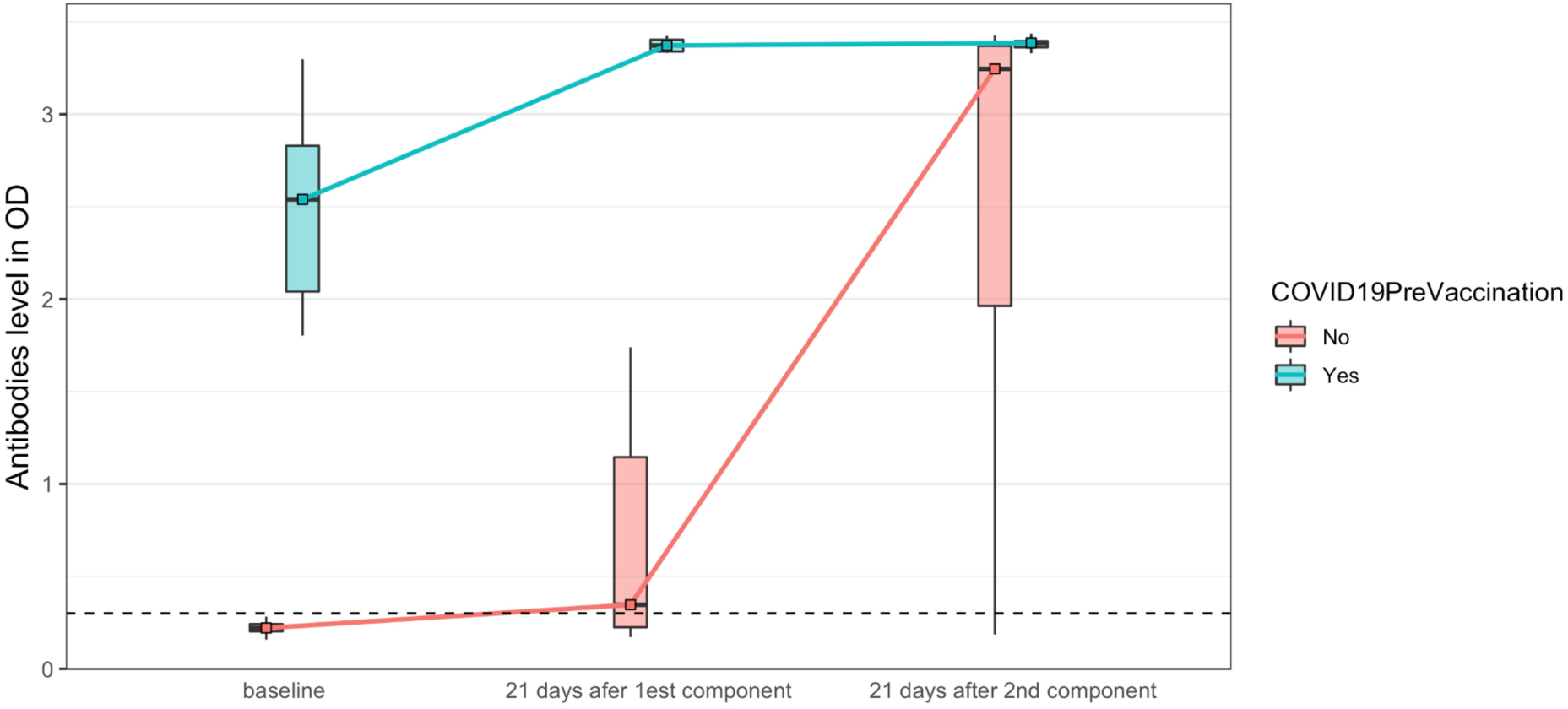
Change in antibodies IgG against SARS-Cov-2 in each measure according of having COVID-19 before vaccination or not Figure shows change in antibodies levels in each measure in patients according to whether they had or not COVID-19 before vaccination. Solid lines join to the median in each box plot. Dash line shows measure under detection limit (no reactive)

## Discussion

This study shows that Sputnik V COVID-19 vaccine in dialysis patients had a low rate of ESAVI and good immunogenicity. Regarding adverse events, none were characterized as a major event, the most frequent ESAVI was pain at the injection site with both components of the vaccine. Considering systemic symptoms, new or worse muscle pain and fever were the most frequent and none of the ESAVI required hospitalization. ESAVI were less frequent compared to phase 2 and 3 studies of the vaccine and also lower compared to the ESAVI in health workers (17). This could be due to the widespread use of antipyretics prior to vaccination. Studies in dialysis patients with BNT162b2 mRNA COVID-19 vaccine showed that also local pain was the most frequent ESAVI in dialysis patients, and diarrhea, fatigue and myalgia among the sistemics (18,19). Symptoms were less frequent in the second component. We found that history of allergy, young age and history of transplantation were associated with a higher frequency of adverse events.

Regarding the seroconversion rate, almost 40% of the patients did not achieve a detectable anti-SARS-Cov-2 IgG antibody titer with the first component, but had a significant increase with the administration of the second component. Patients with COVID-19 prior to vaccination reached almost maximum levels of antibodies at 21 days of the first component, remaining stable at 21 days of the second. Almost 98% of our population had detectable antibodies after vaccination at that time. Hypo-response to vaccines in general has been described in dialysis patients, in vaccination against hepatitis B, which shows a seroconversion of only 40-70% compared to more than 95% in healthy controls (20), attributing associated factors such as age, the presence of diabetes, nutritional status and altered innate and adaptive immune response (21). However, in our experience, the level of seroconversion with the two Sputnik V components was, on the contrary, much higher, and has already been described with other COVID-19 vaccines in dialysis patients (19,21–26), thus highlighting the importance of a complete vaccination in this patients (19,26). Besides, patients who presented COVID-19 after the first Sputnik V component had mild forms of the disease, as has been seen in the general population (27).

Taking into account the group of patients who had detectable anti-SARS-Cov-2 IgG antibodies before the first dose, without a clinical history of COVID-19, thus considered asymptomatic patients, has been already described in the literature (15,28). This group of patients as well as those with known COVID-19 presented a significant seroconversion with the first component (19), this is not reported in most of the other studies since these patients were generally excluded. The presence of SARS-Cov-2 infection was not associated with a higher frequency of ESAVI. Recent studies show that the majority of patients with COVID-19 prior to vaccination develop robust and durable immune responses at 6 months, with less than 5% no evidence of humoral and cellular immunity (29,30). However, preliminary studies carried out with another type of vaccine showed that in the case of those immunized without previous infection, there was a drop in antibody levels at six months, considering the need for a third dose (31,32). In this context, determining which subgroup of dialysis patients would need a booster dose according to their characteristics, comorbidities and type of vaccine received deserves to be investigated in the coming months.

Our work has the strength of being the first published report on the safety and efficacy of the Sputnik V vaccine in dialysis patients, especially considering that this vaccine has not yet been recognized by the WHO. The weakness of this study was not being able to evaluate cellular immunity, two reports (18,24) found a cellular immunity (T response) close to 60% of those vaccinated, less than humoral immunity found in our study which was almost 100%. In any case, the effectiveness of the vaccines implemented in dialysis patients will be demonstrated by the reduction in the infection rate as well as the fatality rate.

Dialysis patients constitute a vulnerable population for SARS-Cov-2 infection, beyond the recommendations that were implemented by dialysis units (33), full vaccination is a priority and necessary. The Sputnik V vaccine has been shown to be safe and effective in this patient population.

## Data Availability

All data produced in the present study are available upon reasonable request to the authors

## Notes

### Competing Interest Statement

The authors have declared no competing interest.

### Author Declarations

Ethics Committee of the Alberto C Taquini Institute for Translational Medicine Research of the Faculty of Medicine of the University of Buenos Aires.

